# Burdwan University Thalassemia Severity (BUTS) Scoring System: A numerical Method For Defining the Clinicopathological status of Thalassaemia Patient

**DOI:** 10.1101/2021.05.01.21256431

**Authors:** Anupam Basu, Prosanto Kumar Chowdhury, Tamoghna Chowdhuy, Sukhendu Sadhukhan, Pabitra Kumar Chakrabarti, Dipankar Saha, Debashis Pal

## Abstract

**Background:** β-haemoglobinopathies are the most common monogenic disorders worldwide. They present with extreme clinical heterogeneity, which makes generalized therapeutic approaches often ineffective. Currently available risk stratification approaches are either too complicated to be deployed at a primary healthcare level or are limited in their applicability across the spectrum of haemoglobinopathies. All current systems also classify patients into the three categories of mild, moderate and severe, where the moderate category is not well-characterized in terms of their expected prognosis.

**Methods:** The current study proposes a severity scoring scheme, utilizing five clinical parameters, viz., steady-state average pre-transfusion haemoglobin, age at presentation, transfusion interval, palpable splenomegaly and growth retardation to classify patients of various β-haemoglobinopathies into severe and non-severe categories. The study then proceeds to validate this scoring scheme on a clinically heterogeneous cohort of 224 Eastern Indian β-thalassaemia and β-haemoglobinopathy patients, and evaluate the predictive performance of the proposed scheme against a clinical standard.

**Results:** All of the chosen parameters except steady-state haemoglobin display strong individual contribution to the final determination of severity, though steady-state haemoglobin conferred increased discriminatory power to the overall scheme. The proposed system achieved an accuracy of 94% against the clinical standard.

**Conclusions:** The proposed risk stratification strategy, being almost entirely empirically-determined, should possess wider applicability across the spectrum of β-haemoglobinopathies than currently existing systems, and should also be more suitable than said systems for studying genotype-phenotype correlations especially within the Eastern Indian β-haemoglobinopathy population.

## INTROUDCTION

Thalassaemia syndromes or Haemoglobinopathies are the most common class of single gene disorders worldwide. Different thalassaemia-causing mutations of the HBB gene have variable impact on the on the quantitative or qualitative production of the globin protein and overall function of the hemoglobin. The genetic variants may be the main force behind the differential clinical presentation or severity of the disease. As per earlier conventions, the severity of the patients, suffering from Thalassemia, has been classified into Major, Intermedia and Minor categories, depending on the frequency of transfusion required for survival [1].

It has been observed that anaemia may not be the sole determinant of thalassaemic disease severity, other factors are also responsible for overall clinical status. Accordingly, clinical conditions of the thalassemia patient, cannot be classified based on the transfusion status. Phadke et al. (2006), categorize disease severity based on multiple parameters [2]. Sripichai et al [3], classified HbE/β-thalassaemia into the 3 categories of mild, moderate, and severe. Another classification was also introduced by Thalassaemia International Federation (TIF) [4], which ‘was bit modification Sripichai et al 2008 [3]. Nonetheless, these three classifications categorize the disease into 3 categories in order of severity.

The TIF guidelines classified the Thalassemia in terms of transfusion requirements [5,6]. Accordingly, clinical conditions may be defined as two categories: Transfusion dependent thalassemia (TDT) or severer and the non transfusion dependent thalassemia (NTDT) or non-severe. This is based on the class of patients who did not need “regular” life-long transfusions for “survival”, though they might need transfusions for a period of time in certain clinical situations such as splenomegaly [5, 6]. Weatherall et al [7] and Musallam et al [8] define the TDT category as corresponding to the severe category and the NTDT category as corresponding to the moderate and mild categories, However, there were no strict and quantitative standalone definitions of “regular” and “survival” provided in the literature. Further, later studies showed the spectrum of symptoms in NTDT individuals to be quite heterogeneous and dynamic, making the physician’s objective to provide minimal and regular intervention with transfusion so as to avert certain organ-specific complications like cardiac remodelling and splenomegaly. Cappellini et al, 2015 [9], proposed a more detailed clinical classification for adult and paediatric NTDT patients, which could dynamically assess individual disease severity. However, a very large number of parameters, and a limited overall scope renders this system unsuitable for most primary physicians. Later, Wiwanitkit and Wiwanitkit criticized the system proposed by Capellini et al as a Letter to the Editor in the same journal [10].

The need was felt for a defined, prognostically-oriented, simple categorization thalassemia in two categories with Severe and non severe with minimal judgmental parameters, for optimal utilization of treatment resources and ensure physiological and social well-being. In this study, our group has attempted, based on a minimal set of clinically relevant parameters, to categorize the severity of individuals across the spectrum of qualitative and quantitative β-globin disorders. Based on the previous experience, we have hypothesized scoring matrix, and this proposed scoring matrix has been validated with validation cohort. Based on this scoring system an android application has also been developed, which is freely available to the android phone.

## MATERIALS AND METHODS

### Design of the Scoring System

For designing an overall severity scoring and classification system, multiple parameters based on previous literature and suggestions by the clinicians were considered. These parameters included steady-state average pre-transfusion haemoglobin, frequency of transfusion, age at first disease presentation (first symptoms of anaemia) or first transfusion, size of palpable spleen (splenomegaly), and the height percentile [11, 12].

The proposed scheme allows a minimum total score of 0 up to a maximum total score of 10.5, with higher scores indicating greater disease severity. The cut-off total score for declaring an individual as severe is decided as 5 or higher. The scoring scheme is presented in Table 1.

**Table 1:**
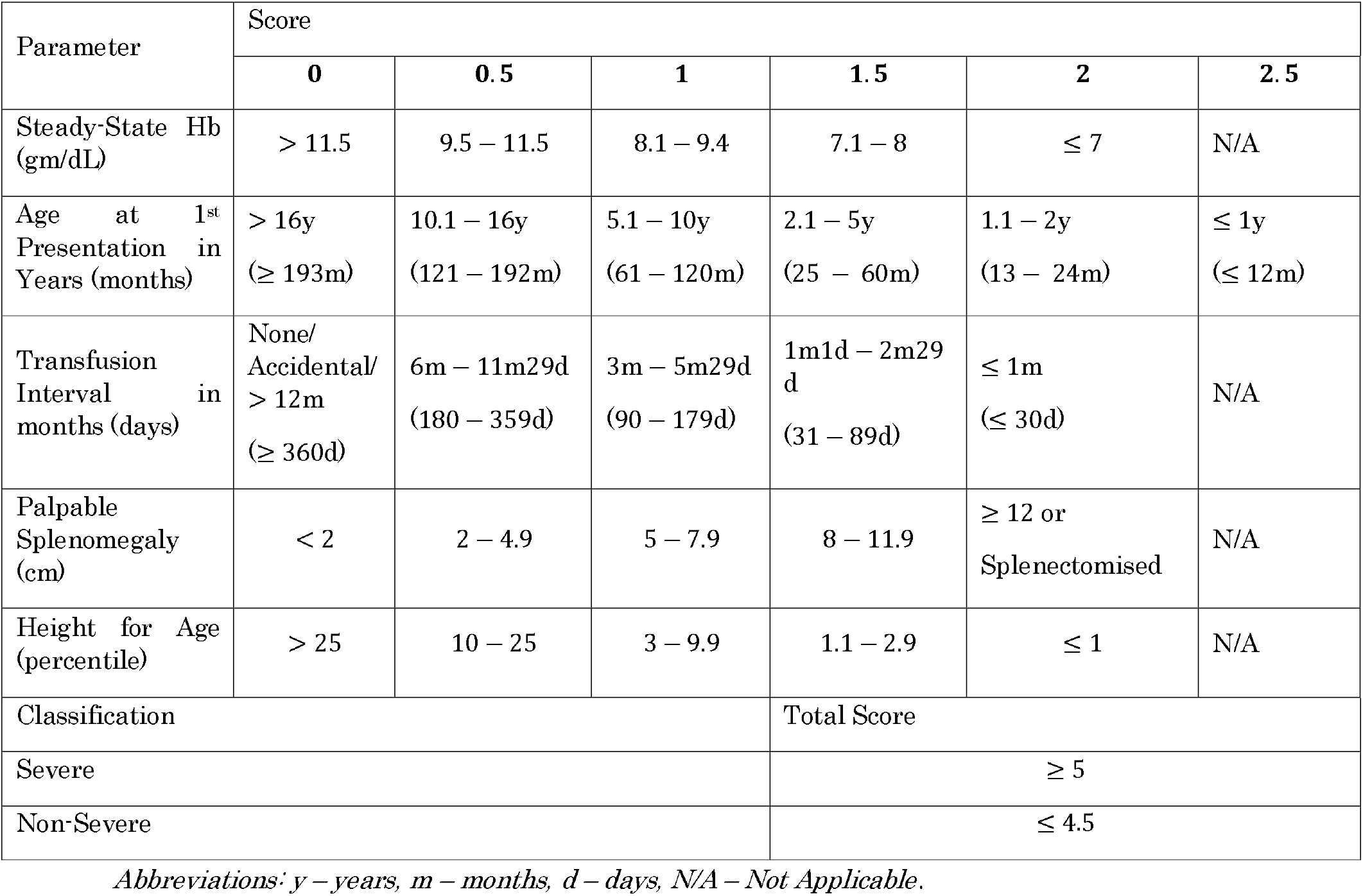
Parameter Scores Assigned with Corresponding Parameter Intervals.

Steady-state haemoglobin, frequency of transfusion, splenomegaly, and height percentile, were scored in between, 0 to 2 in increments of 0.5. Here, 0 implied that the range of the parameter was effectively normal, while 2 implied that the range was severely abnormal. Age at presentation was given more weightage by assigning it an extra interval corresponding to a score of 2.5.

### Validation of Scoring System

#### Subject Recruitment

The current study was based on a cohort of 224 β-thalassaemia and haemoglobin E disease and other haemoglobinopathy patients from the eastern region of India. The hemoglobinopathy status of each patient was confirmed HPLC and Genetic Diagnosis. This study was done as part of a running study to understand the prevalence of different thalassaemia genotypes in Eastern India, approved by the Ethics committee of University of Burdwan (West Bengal, India).

#### Clinical Assessment

Clinical conditions of the patients were evaluated by the one of the authors (PC), who is very experienced in the area of hemoglobinopathy. Anthropometric, biochemical, and haematological data were considered. known complications of congenital haemoglobinopathies like osteopathy, cardiopathy and endocrinopathies were also taken in to considerations. Based on the overall clinical conditions and minimum of 2 years to 12 years follow-up response of the individual patients, they were classified in to sever and non-severe category. Thus, accordingly clinical judgemental classification in to severe and non-severe by this author for the individual subjects were considered as the gold standard for validating the proposed BUTS scoring for the respective subjects and used for the assessing sensitivity and specificity of the proposed system.

### Development Of Android Application for Use of the BUTS scoring system at mobile phone

Newly proposed BUTS scoring system, can be used through Android phone. Accordingly an android application has been developed and can be downloaded from google play store of the Android phone.

### The followings are the input data

1. Steady-State Hb (gm/dL)
2. Age at 1^st^ Presentation
3. Transfusion Interval in months (days)
4. Palpable Splenomegaly/Splenectomised
5. Current Age
6. Current Height

Based on the above input data, application can calculate the severity score and give the output as decision either: Severe state

OR

Non-Severe state

This program can be accessed through the following link: https://play.google.com/store/apps/details?id=org.bu.thalassemiastate

### Statistical Analysis

Pearson correlation analysis was performed to confirm that the parameters considered for the scoring were reasonably independent of each other.

For validating scoring boundaries for individua parameters, a univariate logistic regression analysis with the region of inflection of the regression curve were considered. ROC analysis was performed against the clinical severity classification to validate the choice of cut-off total score for declaring severe status.

## RESULTS

### Independence of Parameters

All parameters under consideration – steady-state pre-transfusion haemoglobin, frequency of transfusion, age at presentation, size of palpable spleen (palpable splenomegaly), and height at current age – were verified to be independent of each other. When the one parameter, compared with other parameter, shows no Pearson correlation (Table 2).

**Table 2:** Pairwise Correlation Matrix of Parameters Considered for Scoring. Numerical Values in the table are pairwise Pearson correlation coefficients or *r* values; |*r*| > 0.6 was considered the threshold for good correlation.

|  | Steady-State Hb (gm/dL) | Age at Presentation (years) | Transfusion Interval (months) | Palpable Splenomegaly (cm) | Height for Age (percentile) |
| --- | --- | --- | --- | --- | --- |
| Steady-State Hb (gm/dL) | **1** | −0.30 | −0.23 | −0.39 | 0.16 |
| Age at Presentation (years) |  | **1** | 0.51 | 0.27 | −0.12 |
| Transfusion Interval (months) |  |  | **1** | 0.14 | 0.05 |
| Palpable Splenomegaly (cm) |  |  |  | **1** | −0.15 |
| Height for Age (percentile) |  |  |  |  | **1** |

### Validation of Scoring Intervals for Chosen Parameters

To validate the choice of the major class boundary of the scoring intervals towards non severe to severe for each parameter, a univariate logistic regression between a parameter and the clinically assessed severity was performed (Figure 1 A to E). For the parameters of steady-state haemoglobin (1A), transfusion interval (1B), age at presentation (1C) and splenomegaly (1D), the region in which the regression curve inflected was within the interval corresponding to a score of 1.5, while being close to the left boundary of these intervals with the intervals corresponding to scores of 1. In case of height percentile (1E), the regression curve inflected in a region within the interval corresponding to a score of 0.5, though it was close to the right boundary of this interval with the interval corresponding to a score of 1. This confirmed that the choice of the intervals for middling severity (corresponding to scores of 1), as it was around this interval that the correspondence of parameter values with the clinical severity changed most abruptly. Though the assignment of the other intervals was unable to be directly verified, it can be seen that in all cases, the choice of the intermediate interval boundaries lays within the inflection region instead of the asymptote region of the regression curve, indicating changing probabilities for being clinically severe as the parameter value changes.

**Figure 1:**
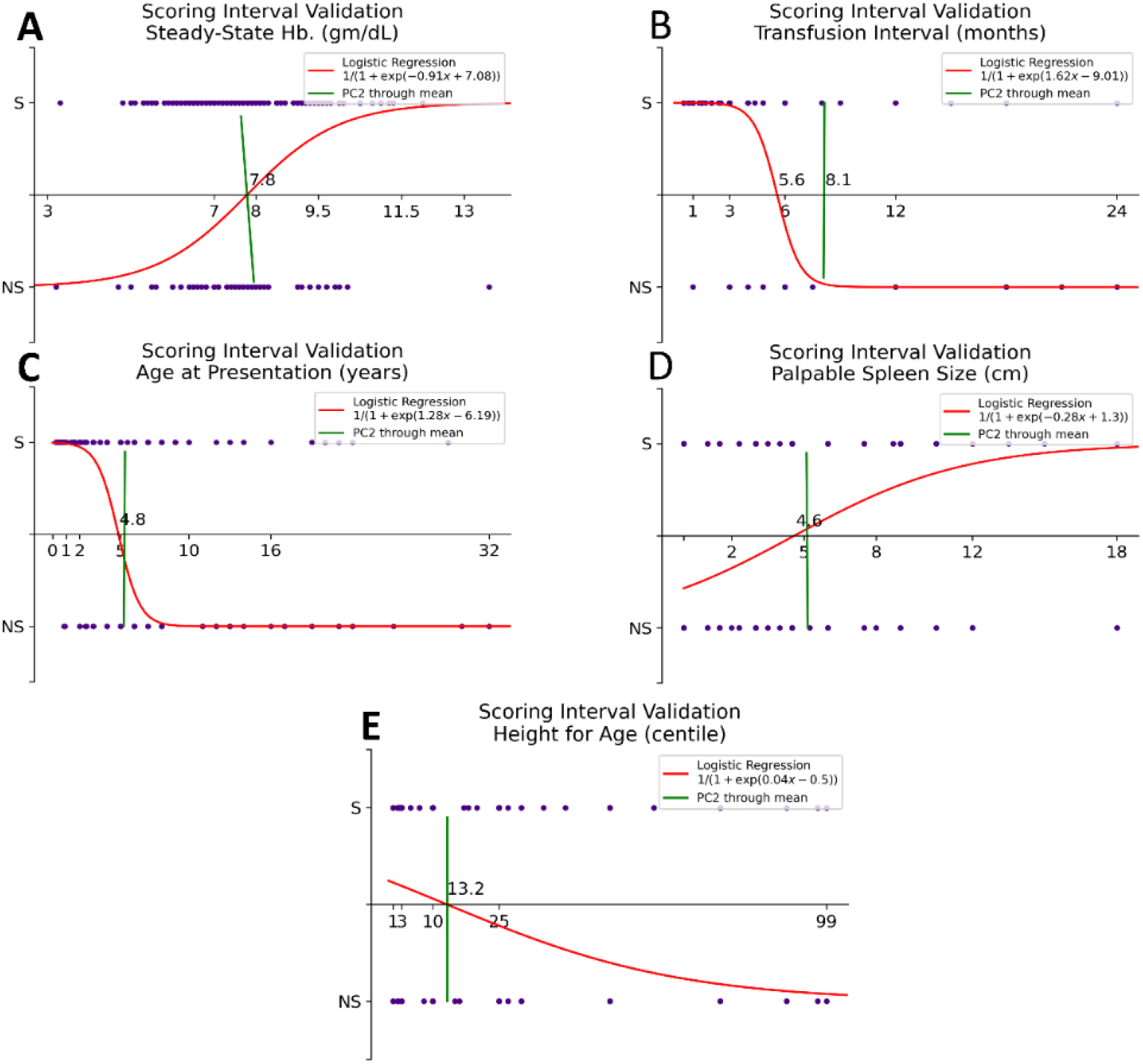
Validation of Choice of Scoring Intervals for Parameters. Univariate Logistic Regression (red) was performed to relate each scored parameter, *i.e*., (A) – Steady-State Hb, (B) – Transfusion Interval, (C) – Age at Presentation, (D) – Palpable Spleen Size, and (E) – Height for Age, to the clinical severity. The Y-axis represents the probability of an individual being severe, with the X-axis representing the range of the parameter (ticks are at scoring interval boundaries). Principal Component 2 through the mean of the data (green) is also indicated in each plot. Inflection points for (A) through (D) correspond to a score of 1.5 but are close to the interval limit for a score of 1, while the inflection point for (E) corresponds to a score of 0.5 but is close to the interval limit for a score of 1.

### Validation of Cut-off Score

To verify the choice of a total score of at least 5 as the cut-off for classifying an individual as severe, a Receiver Operating Characteristic (ROC) curve analysis was performed (Figure 2). As seen in Figure 2, we determined that the true positive rate was maximized and the false positive rate was minimized, and thus the best categorical segregation was indeed afforded, by the choice of a cut-off of 5 out of 10.5.

**Figure 2:**
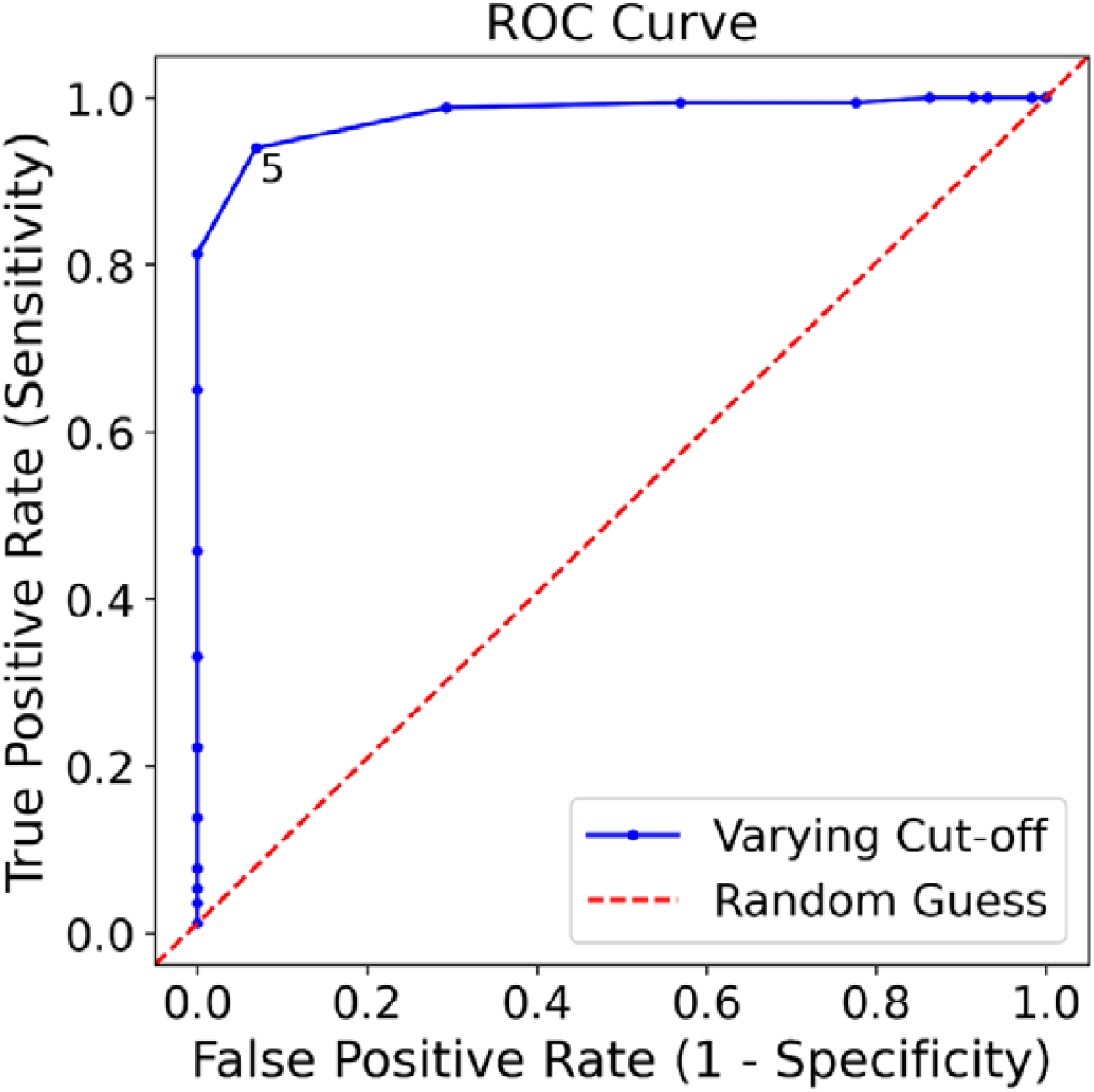
Receiver Operating Characteristic (ROC) curve for determination optimal cut-off score.

**Figure 3:**
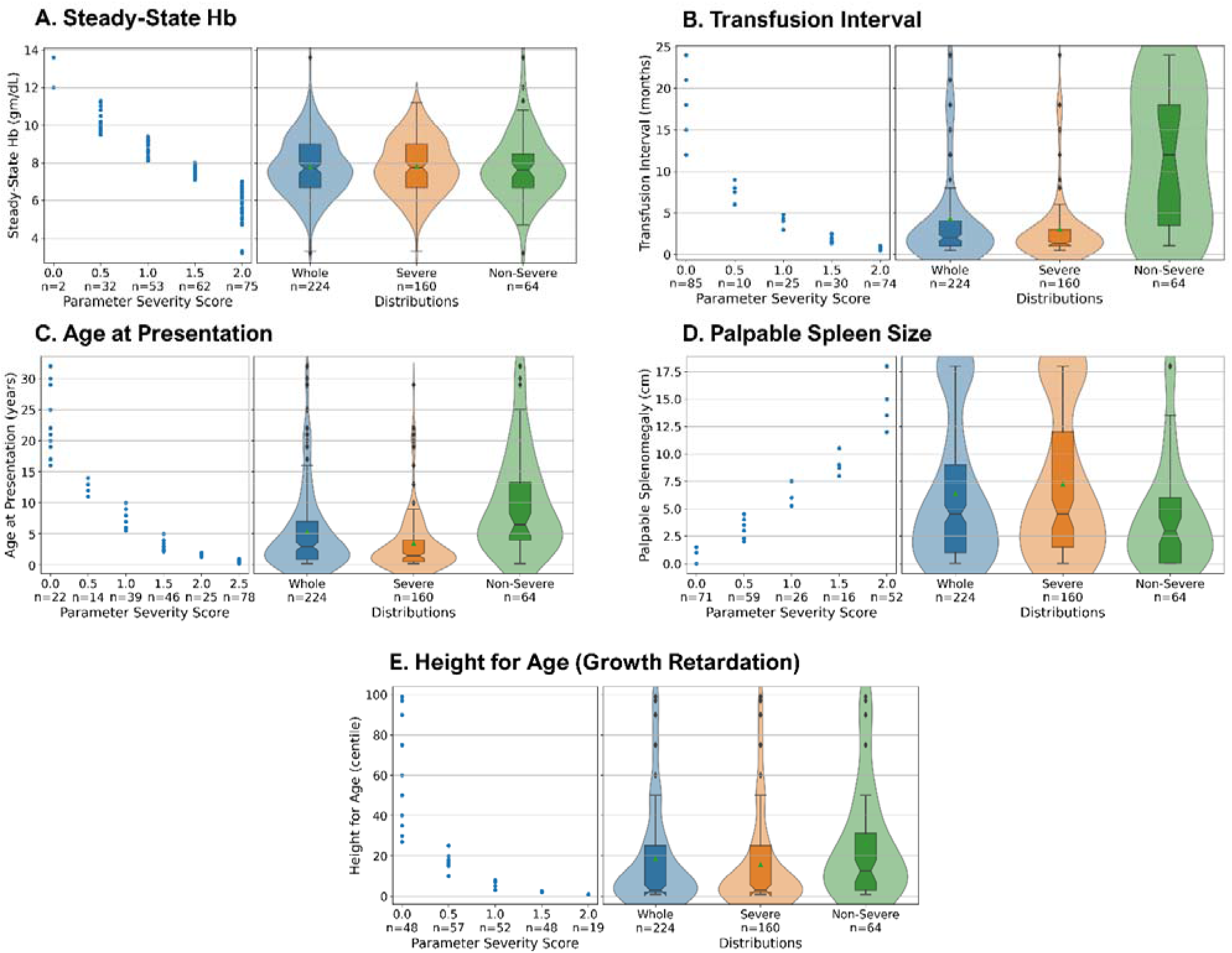
Box-whisker and violin plot for validation class boundaries of the individual parameters against severity classes. The range and frequency distributions of the parameters are plotted as box-whisker plots superimposed on a violin plot, respectively.(A) Steady-State Haemoglobin (higher implies less abnormal), (B) Transfusion Interval (higher implies less abnormal), (C) Age at Presentation (higher implies less abnormal), (D) Palpable Spleen Size (lower implies less abnormal), (E) Height for Age (higher implies less abnormal).Note that in (D), patients who have been splenectomised are represented by the maximum observed spleen size of 18cm, and in the Right Panel the bimodal distribution in the case of the overall and severe categories reflect the much larger fraction of patients who have undergone splenectomies (22.5%) as compared to the non-severe category (6.2%).

The True Positive Rate or Sensitivity is plotted against the False Positive Rate (1 – Specificity) as the discrimination threshold (here, the cut-off score for declaring an individual as severe) is varied. Each individual cut-off score from 0 to 10.5 in steps of 0.5 corresponds to a blue point along the curve. Sensitivity and Specificity are simultaneously maximized at a cut-off score of 5 out of 10.5, and the steepness of the curve indicates that classifications are accurate only in a narrow range of cut-off scores around 5.

### Performance comparison of present Scoring System with actual clinical judgment

The proposed scoring system was then compared with the clinical judgement. As per present scoring system (BUTS), out of 224 patients, 64 have been appeared as non-severe category and 160 have been appeared as severe category. Based on the clinical judgment, 58 have been appeared as non-severe and 166 appeared as severe (Supplementary data table S1). The clinical judgment was done by one of the experienced authors based on different clinical parameters along with parameters considered for making the scoring system. The other parameters, which were considered for the clinical judgement – weight, BMI, Growth retardation, leg ulcers, EMH, Genu Valgum, Cranio-facial deformity, DCM, Endocrine and Sexual Immaturity, Liver Fibrosis/cirrhosis, Thrombosis and others. In comparison with clinical judgement the sensitivity and specificity were 93.9% and 93.1% respectively (Table 3). On the other hand, positive predictive value (PPV) and negative predictive value (NPV) were 97.5% and 84.37% respectively (Table 3).

**Table 3:** Sensitivity, specificity and positive and negative Predictive Performance BUTS scoring system.

|  |  | Clinical Judgment |  |
| --- | --- | --- | --- |
|  |  | Severe (166) | Non-Severe (58) |
| **BUTS Scoring System:** | Severe (160) | True Positive (a): 156 | False Positive (b): 4 |
|  | Non-Severe (64) | False Negative (c) : (10) | True Negative (d): (54) |
| **Sensitivity:**<br>=$[a/(a+c)] \times 100$ | 93.97% | | |
| **Specificity**<br>=$[d/(b+d)] \times 100$ | 93.1% | | |
| **Positive predictive value (PPV)**<br>=$[a/(a+b)] \times 100$ | 97.5% | | |
| **Negative predictive value (NPV)**<br>=$[d/(c+d)] \times 100$ | 84.37% | | |

## DISCUSSION

From the time of identification and description of the first case of non-spherocytic haemolytic anaemia, by Drs Thomas Cooley and Pearl Lee, the diagnosis and pathophysiology of the so-called Cooley’s Anaemia alias Thalassaemia has come a long way. This has been the model disorder as regards molecular biology and translational biochemistry. Different genetic loci have been identified and their roles in the pathophysiology described. To our surprise what was a mystery then is still a mystery now. For decades, 1925 till date, clinicians and scientists alike, have tried to classify Thalassaemia, rather Thalassaemia Syndrome, according to clinical severity, because it became very apparent since the first description of the disorder – though looked alike microscopically alike, did not behave the same in all suffering from it. It was a heterogenous group, some requiring frequent blood transfusions to be kept alive, while, few maintained themselves to adulthood even old age, were clinically normal or compensated not requiring any or little transfusion support. Blood was and still is, a scarce commodity, a classification was felt required to rationalize the treatment and also justify the appropriate usage of the precious liquid tissue. Initially, those requiring frequent transfusions were classified as Major and less transfusions as Intermedia and no transfusions as Minor. Later it was realized that the Minors were actually carriers or heterozygous for the defective gene responsible for causing thalassaemia. As, this was the disease of low and middle socio-economic strata, where malnutrition and intercurrent infections were very common, plenty of children presented with very low haemoglobin at a very early age, thus were blindly started on regular blood transfusion regime, who were later learned to produce enough usable haemoglobin, to survive till adulthood with little or no transfusions at all. This misdiagnosis of disease severity not only led to squandering of the precious blood and also causing functional harm to the body’s vital organs due to accumulation of excess iron. Till date, five attempts have been made by our learned teachers to classify this disorder, so as to ensure justified utilization of resources and ensuring socially and economically productive lives of the ones optimally managed. There are basically two targets which require to be fulfilled, while classifying this disorder:

1. The classification should correctly categorize the severity of the disease at first or latest by the third contact visit with the doctor.
2. The classification should be based on very simple clinically elicitable and biochemically feasible, easy and cheap, procedures and tests.
3. It should classify the disease into two broad categories with acceptable certainty, not to accommodate a third group in between two extremes, whose treatment modality will be ambiguous.
4. It should be able to classify all the different disorders clustered under thalassaemia syndrome – one size fit all

After studying the classifications that are in vogue till date, it was realized that no one was satisfying all the criteria given above. So we, chose from our group of patients suffering from thalassaemia syndromes, who were followed up form a minimum of 5 to a maximum of 18 years, to get an inkling about the disease’s individualistic natural history.

For our study, the parameters that were chosen for categorizing the severity *viz:* steady-state haemoglobin, transfusion interval, age at first presentation or transfusion, splenomegaly, and height percentile, to be the minimal set of criteria which are directly instrumental for the phenotype clinical state of the patient. The information about these parameters is easily possible to get for low economic back ground patents also. Thus, assessment based on these parameters could be very helpful for the treating physician.

The choice of scoring intervals for each parameter were based on physiological considerations, of the patients as per clinical experience. Accordingly, for steady-state haemoglobin, the extreme scoring intervals (0 and 2) were chosen based on physiological extremes. Average haemoglobin of ≤ 8 gm/dL is often associated with cardiopathies, especially iron overload cardiomyopathies in regularly transfused thalassaemic individuals [13, 14], while average haemoglobin above 11.5 gm/dL was considered effectively normal by the clinicians. The intermediate intervals were chosen to have an approximately equal number of individuals in each interval. For growth retardation as measured by the height for age as percentile of population. the scoring intervals were chosen based on Indian Academy of Paediatrics guidelines, based on WHO, for judging growth retardation and malnutrition [11]; growth above the 25^th^ percentile is considered normal while growth below the 3^rd^ percentile represents severe growth retardation. Growth near the 10^th^ percentile is often considered the threshold for clinically significant growth retardation [12]. In this scoring system, w eight for age was not considered as a parameter, as the clinicians believed that weight could fluctuate rapidly for reasons unrelated to the disease being studied - nutritional status, bladder, and bowel habits, etc, which may make the individual lose the gained weight. Similar fluctuations do not happen with height; the rate of height gain may be compromised or even made static, but height could not be lost and thus complicate cumulative assessments.

Organomegaly was considered by judging the size of the palpable spleen. The size was taken after palpating the spleen and measured in cm or fingers (1 finger = 1.5 cm) below the left costal margin along the splenic axis. The scoring intervals were chosen to be regular and to have an approximately equal number of individuals in each interval, with non-existent or mild splenomegaly (<2 cm) being considered effectively normal. Splenectomised individuals were considered maximally severe. Liver palpability was not considered as it could vary with other conditions like malaria, typhoid, and/or heart failure, which might not be related to the disease under study.

Age at presentation was the earlier of the age at which anaemia was first diagnosed and the age at which the first transfusion was received (if at all), this flexibility being allowed to compensate for incomplete medical records. The clinicians unanimously considered any individual presenting at 2 years of age of lower to be severe, and this parameter was considered by them as the most important parameter for defining the disease severity. As such, a higher scoring weightage was given to children presenting at or below 1 year of age. An upper limit of 16 years was chosen as the point at which an individual could be considered physiologically adult, while the intermediate cut-offs of 5 and 10 years were chosen to have a roughly equal number of individuals in each sub-interval.

Transfusion interval, is also a very important parameter for defining the severity state of the patient. It has been observed that most of the severe patient needs transfusion within 30 days interval. Thus, highest severity score has been given for the patient needing transfusion within 30 days. A yearly or less frequent transfusion has been considered as less severe state of the disease, accordingly given very low severity scoring. The intermediate intervals were chosen to have an approximately equal number of individuals in each interval. The lowest scored category corresponds to yearly or less frequent transfusions, encompassing a large range which was considered by the clinicians to indicate transfusion for reasons unrelated to the chronic anaemia.

Retrospective classification of the disease severity of patients with thalassaemia and similar disorders provides little more than academic utility, as the patient is usually no longer in a position to significantly benefit from any modified therapeutic approaches. For the patient to have a higher quality of life with optimal utilization of available resources, a criterion to effectively triage the patient on the basis of their disease severity and the expected extent of therapeutic intervention necessary, is required as soon as possible after the initial presentation. This would provide the attending physician with the basic information necessary to decide on an individualized course of therapy.

Currently existing disease severity stratification systems for thalassaemias and related disorders tend to focus on limited genetic subsets of the overall spectrum thalassaemia syndromes. The system recommended by the Thalassaemia International Federation [4], based on the scoring system originally proposed by Sripichai et al in 2008 [3], is limited to considering only β-thalassaemia/HbE disease populations. This could be considered a significant shortcoming of current systems, as most thalassaemias present with a similar set of superficial symptoms (haemolytic anaemia and associated complications) and thus should be able to be classified on a purely phenotypic basis without limiting the genotypic variability of the studied cohort. However, clinically oriented scoring systems like the one proposed by Capellini et al in 2016 [9] suffer from an alternate problem, in that they are usually quite complicated for primary practitioners to utilize as a first response measure.

The developed BUTS system classifies patients of common β-thalassaemia and β-haemoglobinopathy syndromes in to severe and non severe category, those are HPLC confirmed status. The proposed system forgoes the inclusion of simple heterozygotes (carriers) and also does not attempt to correlate itself (currently) with the complex intergenic interactions which often characterize thalassaemia. We have accepted the phenotype and tried to classify the phenotypic severity so as to formulate the most suitable therapeutic intervention.

Previous studies such as Sripichai et al [3] have performed grounds-up investigations to determine the strongest contributing factors to phenotypic severity in thalassaemic cohorts. Though the final set of scored parameters proposed therein are viable to be employed in a primary clinical setting, the process and reasoning for the choice of these parameters is considerably opaque to most clinicians. The proposed system posits the scoring scheme *a priori*, based on physiological and clinical considerations first and foremost, which we then proceeded to validate in a clinically heterogeneous thalassaemic cohort. We have also not classified an intermediate category, as an intermediate category corresponds to a survival “grey zone”, which is not useful to the physician for deciding the basic course of treatment. But on the same time scoring value can guide the clinician about the degree of severity or non severity.

The criterion, that patients could only be categorized as either Severe or Non-Severe based on the overall clinical status. Patients who require significant medical and/or surgical interventions to keep them alive and productive till attainment of adulthood or sexual maturity are severe, while those who do not require such significant interventions are non-severe. We considered five clinical and biochemical parameters for quantifying and classifying the disease severity, which were chosen based on prior literature as well as clinical considerations. We expect that this empirical basis for our scoring scheme would render it less biased as compared to solutions devised from the ground-up using data-driven approaches.

Further contention arises from the fact that existing literature considers the Non-Transfusion-Dependent Thalassaemic (NTDT) category to correspond to the mild and moderate categories established in the Mahidol scoring system established by Sripichai et al and Taher et al [3, 4], though for enabling comparison of the existing systems to our proposed system, we had to consider the severe and moderate categories of the Mahidol system [3] as severe, and their mild category as non-severe. Along with the demonstrated increased specificity and positive predictive value of the proposed system, this also illustrates the somewhat ambiguous nature of the currently existing classification systems as regards prognostic impact. The currently proposed system is considerably less ambiguous in terms of prognostic impact, while having a much more well-defined non-severe category in terms of age of presentation and transfusion interval.

The proposed system iteratively improves on the mentioned shortcomings of the currently existing systems for classifying thalassaemia disease severity and should have demonstrably increased clinical utility compared to the prior systems. The current system is organized with a strict clinical basis, and individuals can be classified for severity as early as three received transfusions, at less than 1.5 years of age. The scoring system can also be packaged and provided as an online utility or application for tracking patient data, severity and treatment strategies and outcomes as part of a holistic treatment assessment platform for clinicians. At present, an online tool has been developed to classify individual patient severity using this system. In the future, assessments of the impacts of genotypic determinants on phenotypic severity could also be performed against this proposed system, cementing the clinical impact of certain genetic variations in case of thalassaemia.

This scoring system, can be applied at the stage of first contact and even then it will be able to reasonably classify the disorder according to severity, rather than waiting for multiple visits, followed by complex biochemical, radiological and imaging investigations for the more accurate diagnosis of severity, because this delay in institution of proper management, due to which the complications have developed, an accurate retrospective classification of severity would be a futile attempt to clinically bring the individual under question back on optimum clinical track. This new classification system, when applied, will enable even the semi-trained rural practitioner to correctly attribute the severity of the disorder utilizing the clinical acumen and minimal biochemical test and rationally allocate precious resources with preferential and individualized management strategy, to ensure, socially and economically productive individuals, in resource restricted settings.

## Data Availability

If the raw data is needed it will be provdied on request with proper procedure

## Funding information and Acknowledgments

Authors are thankful to DST SERB, FIST and PURSE support. The authors are also thankful to the Department of Biotechnology, Govt. of India for supporting this work [Sanc. No-BT/PR26461/MED/12/821/2018].

