## Supplementary material for "Burdwan University Thalassemia Severity (BUTS) Scoring System: A numerical Method For Defining the Clinicopathological status of Thalassaemia Patient": supplimentary table

Table S1: Severity Status of the 224 subjects, as per clinical judgment and also by BUTS scoring method [S = severe, NS = non severe ]

| **Sl. No.** | **Gender** | **Clinical judgment** | **BUTS Score** |
| --- | --- | --- | --- |
| **1** | M | S | 7.5 |
| **2** | M | NS | 3 |
| **3** | M | NS | 3.5 |
| **4** | M | NS | 3 |
| **5** | F | S | 5.5 |
| **6** | F | NS | 4 |
| **7** | M | S | 6.5 |
| **8** | F | S | 7 |
| **9** | F | NS | 4 |
| **10** | F | S | 6 |
| **11** | M | S | 5 |
| **12** | M | S | 8 |
| **13** | M | S | 5 |
| **14** | F | S | 5 |
| **15** | M | S | 7 |
| **16** | M | S | 5 |
| **17** | M | S | 5 |
| **18** | M | NS | 5 |
| **19** | M | S | 6.5 |
| **20** | M | NS | 4 |
| **21** | M | S | 5 |
| **22** | M | S | 8 |
| **23** | M | NS | 4.5 |
| **24** | F | NS | 4 |
| **25** | M | S | 5.5 |
| **26** | M | NS | 3.5 |
| **27** | M | S | 6 |
| **28** | F | S | 8.5 |
| **29** | F | S | 7.5 |
| **30** | F | S | 5.5 |
| **31** | F | S | 4.5 |
| **32** | F | S | 5 |
| **33** | M | S | 6 |
| **34** | F | S | 5.5 |
| **35** | M | S | 8 |
| **36** | M | S | 7 |
| **37** | M | S | 4.5 |
| **38** | M | S | 4 |
| **39** | M | S | 6 |
| **40** | M | NS | 4 |
| **41** | M | S | 7 |
| **42** | M | S | 5 |
| **43** | M | NS | 4.5 |
| **44** | M | S | 5.5 |
| **45** | M | S | 7 |
| **46** | M | S | 6 |
| **47** | M | NS | 3.5 |
| **48** | F | S | 6 |
| **49** | M | S | 5.5 |
| **50** | M | S | 5.5 |
| **51** | M | NS | 4.5 |
| **52** | M | S | 6.5 |
| **53** | F | S | 5.5 |
| **54** | F | NS | 4.5 |
| **55** | F | S | 7.5 |
| **56** | M | S | 5.5 |
| **57** | M | S | 7.5 |
| **58** | M | S | 5 |
| **59** | M | S | 5.5 |
| **60** | F | S | 6 |
| **61** | F | NS | 4 |
| **62** | M | NS | 3.5 |
| **63** | M | S | 7.5 |
| **64** | M | S | 10 |
| **65** | M | S | 5 |
| **66** | F | NS | 4 |
| **67** | F | S | 6 |
| **68** | F | NS | 4.5 |
| **69** | F | S | 5.5 |
| **70** | F | S | 6.5 |
| **71** | F | S | 5.5 |
| **72** | M | S | 8 |
| **73** | F | NS | 5 |
| **74** | F | S | 6.5 |
| **75** | F | S | 9 |
| **76** | F | NS | 3.5 |
| **77** | M | S | 5.5 |
| **78** | M | S | 9 |
| **79** | M | S | 6 |
| **80** | M | NS | 3.5 |
| **81** | M | S | 7 |
| **82** | F | S | 9.5 |
| **83** | M | S | 8.5 |
| **84** | F | S | 6 |
| **85** | F | S | 7 |
| **86** | M | S | 6.5 |
| **87** | F | S | 6 |
| **88** | M | S | 7 |
| **89** | M | S | 6.5 |
| **90** | F | S | 5 |
| **91** | F | S | 8 |
| **92** | M | NS | 5 |
| **93** | M | S | 6.5 |
| **94** | M | S | 6.5 |
| **95** | F | S | 4.5 |
| **96** | M | S | 7 |
| **97** | M | S | 4.5 |
| **98** | F | S | 6 |
| **99** | F | S | 6.5 |
| **100** | F | S | 5 |
| **101** | M | S | 8.5 |
| **102** | F | S | 7.5 |
| **103** | M | NS | 4 |
| **104** | M | S | 6.5 |
| **105** | M | S | 8 |
| **106** | F | S | 5.5 |
| **107** | F | S | 5.5 |
| **108** | M | S | 5.5 |
| **109** | F | NS | 4 |
| **110** | M | S | 6.5 |
| **111** | M | S | 6 |
| **112** | M | S | 10 |
| **113** | M | NS | 3 |
| **114** | M | S | 6.5 |
| **115** | M | S | 6 |
| **116** | F | S | 6 |
| **117** | M | S | 6 |
| **118** | F | S | 6.5 |
| **119** | M | S | 8 |
| **120** | F | S | 7.5 |
| **121** | M | S | 4.5 |
| **122** | F | NS | 1.5 |
| **123** | M | S | 6.5 |
| **124** | F | S | 6 |
| **125** | F | S | 5 |
| **126** | M | NS | 4.5 |
| **127** | M | NS | 2.5 |
| **128** | F | S | 7 |
| **129** | F | S | 5.5 |
| **130** | M | NS | 4 |
| **131** | M | S | 5 |
| **132** | F | S | 6 |
| **133** | M | S | 7 |
| **134** | M | S | 7 |
| **135** | M | NS | 4 |
| **136** | M | NS | 3.5 |
| **137** | F | S | 8 |
| **138** | M | S | 6 |
| **139** | M | NS | 4.5 |
| **140** | M | S | 5.5 |
| **141** | F | NS | 3.5 |
| **142** | M | S | 6 |
| **143** | M | S | 7 |
| **144** | M | S | 5.5 |
| **145** | M | NS | 4 |
| **146** | M | S | 6 |
| **147** | M | S | 6 |
| **148** | M | NS | 0.5 |
| **149** | F | NS | 1.5 |
| **150** | F | S | 9.5 |
| **151** | F | NS | 4 |
| **152** | F | S | 5.5 |
| **153** | M | S | 6 |
| **154** | F | S | 6 |
| **155** | M | S | 5.5 |
| **156** | F | S | 7 |
| **157** | F | S | 7.5 |
| **158** | M | S | 7.5 |
| **159** | M | S | 6 |
| **160** | M | S | 5.5 |
| **161** | M | NS | 4.5 |
| **162** | F | S | 9.5 |
| **163** | M | S | 5 |
| **164** | M | NS | 5 |
| **165** | F | NS | 4.5 |
| **166** | F | NS | 2 |
| **167** | F | S | 7 |
| **168** | M | S | 7 |
| **169** | M | S | 7.5 |
| **170** | M | S | 4.5 |
| **171** | F | S | 5.5 |
| **172** | M | S | 6 |
| **173** | M | NS | 4 |
| **174** | M | NS | 4.5 |
| **175** | M | NS | 3.5 |
| **176** | M | S | 8.5 |
| **177** | M | S | 6.5 |
| **178** | F | S | 5 |
| **179** | F | S | 7 |
| **180** | M | NS | 4 |
| **181** | F | S | 6 |
| **182** | M | NS | 1.5 |
| **183** | F | S | 6 |
| **184** | M | NS | 2.5 |
| **185** | M | NS | 3.5 |
| **186** | F | S | 7.5 |
| **187** | F | S | 4.5 |
| **188** | F | S | 5.5 |
| **189** | F | S | 7.5 |
| **190** | M | S | 6 |
| **191** | F | S | 5 |
| **192** | F | S | 6.5 |
| **193** | F | S | 7.5 |
| **194** | M | S | 5 |
| **195** | M | S | 5.5 |
| **196** | F | S | 6.5 |
| **197** | M | NS | 3 |
| **198** | M | S | 6.5 |
| **199** | M | NS | 3.5 |
| **200** | M | NS | 4 |
| **201** | F | S | 5 |
| **202** | F | S | 5.5 |
| **203** | F | S | 7 |
| **204** | M | S | 8 |
| **205** | M | S | 6.5 |
| **206** | F | S | 6 |
| **207** | M | S | 9.5 |
| **208** | M | S | 5.5 |
| **209** | F | S | 6.5 |
| **210** | M | S | 6 |
| **211** | M | NS | 4.5 |
| **212** | M | S | 6 |
| **213** | M | S | 8 |
| **214** | M | S | 5 |
| **215** | M | NS | 2.5 |
| **216** | F | S | 5 |
| **217** | F | S | 4.5 |
| **218** | F | S | 9 |
| **219** | M | NS | 3.5 |
| **220** | M | NS | 4.5 |
| **221** | M | NS | 4.5 |
| **222** | F | S | 7.5 |
| **223** | F | S | 3 |
| **224** | F | NS | 3 |
